# Reduced contrast surround suppression associated with schizophrenia depends on visual acuity and scene context

**DOI:** 10.1101/2022.05.10.22273873

**Authors:** Victor J. Pokorny, Michael-Paul Schallmo, Scott R. Sponheim, Cheryl Olman

## Abstract

Perceptual distortions are core features of psychosis. Weakened surround suppression has been proposed as a neural mechanism of such atypical perceptual experiences. While previous work has measured suppression by asking participants to report the perceived contrast of a low-contrast target surrounded by a high-contrast surround, it is possible to modulate perceived contrast solely by manipulating the orientation of a matched-contrast center and surround. Removing the bottom-up segmentation cue of contrast difference and isolating the orientation-dependent suppression may clarify the neural processes responsible for atypical surround suppression in psychosis. We examined surround suppression across a spectrum of psychotic psychopathology including people with schizophrenia (PSZ; N=31) and bipolar disorder (PBD; N=29), first-degree biological relatives of these patient groups (PBDrel, PSZrel; N=28, N=21, respectively), and healthy controls (N=29). Surround suppression deficits in PSZ, while observable under many stimulus conditions, were absent under the condition that produced the strongest suppression. PBD and PSZrel exhibited intermediate suppression, while PBDrel performed most similarly to controls. Intriguingly, group differences in surround suppression magnitude were moderated by visual acuity. We propose a potential model by which visual acuity and/or focal attention interact with untuned gain control that reproduces the observed pattern of results including the lack of group differences when orientation of center and surround are the same. Our findings further elucidate perceptual mechanisms of impaired center-surround processing in psychosis and provide insights into the effects of visual acuity on orientation-dependent suppression in PSZ.

## Introduction

Perceptual distortions are a primary symptom of psychosis. In 2005, Dakin et al.^1^ reported a striking reduction of contrast surround suppression in patients with schizophrenia (PSZ), which has been borne out in several other reports^2–8^. The magnitude of the reduction appears to fluctuate with symptom severity^9^, with more recent studies in out-patient populations estimating smaller effect sizes than reported for the in-patient sample of the 2005 study. Still, the task is valuable because it quantifies the function of well-understood neural mechanisms in primary visual cortex within patient populations. Also, because perceptions reported during the task by PSZ more closely match the physical reality of stimuli, concerns about generalized cognitive deficits impairing performance are diminished.

Several neural mechanisms – occurring both inside (i.e., intrinsic) and outside (i.e., extrinsic) of primary visual cortex (V1) work together to determine perceived contrast, which can generally be predicted from firing rates of neurons in V1^10^. One such mechanism is V1-intrinsic untuned gain control which provides orientation-insensitive suppression that is stronger for more intense (i.e., higher contrast) stimuli^11,12^. Untuned gain control is thought to be weaker in PSZ^13^. In addition to untuned gain control, orientation-dependent mechanisms suppress surrounds that are parallel (or near parallel) to the center while surrounds orthogonal (or near orthogonal) to the center produce little to no suppression ^12,14,15^. There is some evidence that the efficacy of these mechanisms differs for PSZ^16^, though also see ^7^.

Feedback from higher areas in the visual cortex is also known to alter firing rates of neurons in primary visual cortex^14, 17^. In particular, electrocorticography measurements in human V2 and V3^18^ and primate electrophysiology measurements in V1^19^ provide evidence that V1-extrinsic segmentation cues (i.e., object boundaries) modulate V1-intrinsic suppression mechanisms because suppression for parallel surrounds occurs within 50 ms of stimulus onset^20^; however, when a boundary is present, neural responses occurring more than 100 ms after stimulus onset have the same amplitude for both parallel and orthogonal surrounds^18^. This suggests a later abolishment of earlier suppression induced by the parallel surround when a boundary is present. Such an effect is thought to be mediated via feedback, possibly from border-ownership processes in V2^21^ or V4^22^ and may be subject to regulation by attention or awareness. V1-extrinsic mechanisms are likely important to consider in the context of psychosis due to well-documented attentional deficits and evidence of altered top-down regulation of low-level inputs^23–28^.

Typically, contrast surround suppression is measured with a low-contrast target embedded in a high-contrast surround, but in the present study, the luminance contrasts of the center and surround gratings were matched. Although this choice reduces the expected magnitude of the behavioral effect^29^, it also controls for the bottom-up contrast-difference cues that help draw spatial attention and thus is useful for clarifying whether deficits in PSZ are driven by altered attentional or low-level visual processes^30^.

The contribution of V1 intrinsic and extrinsic mechanisms to atypical surround suppression in psychosis can be further clarified by manipulating the distance between the surround and center. The surrounding receptive field can be disaggregated into two regions (termed near and far surround) that are mediated by different neural mechanisms. The far surround is mediated by feedback connections extrinsic to V1 while the near-surround is mediated by a combination of both feedback connections and V1-intrinsic horizontal connections^14,31^. Thus, by assessing contrast surround suppression for both near and far surrounds, we aimed to further distinguish the possible neural mechanisms by which PSZ experience weakened surround suppression.

In earlier work, PSZ have been measured to have a ∼50% reduction in perceptual suppression relative to controls for a task in which a low-contrast drifting center grating was embedded in a high-contrast surround grating drifting in the same direction with no explicit boundaries between center and surround^7,32^. The introduction of additional, explicit boundaries (a gap, a direction difference, or an orientation change) reduced surround suppression for all groups by comparable amounts^7^. Additionally, reduced ability to attend to the stimuli, as measured by catch trials, has also been shown to contribute to the magnitude of the contrast surround suppression^33^. Thus, reduced contrast surround suppression in PSZ appears to be caused by a combination of V1-intrinsic mechanisms (orientation-insensitive gain control and/or orientation-selective mechanisms) and altered spatial attention, but not an alteration of boundary-related facilitation mechanisms. Previous work has not delineated the relative contributions of the two putative V1-intrinsic mechanisms – untuned gain control and orientation-dependent suppression – to perceptual contrast surround suppression, which was the aim of the current work.

It is unclear whether atypical orientation-dependent surround suppression is present in other psychotic disorders such as Bipolar disorder^1,7,34^. Given the criticisms of reliability and validity of categorical DSM diagnoses and the shared features of schizophrenia and bipolar disorder^35,36^, understanding the degree to which visual processing impairments are reflective of categorical differences between disorders or are reflective of a unified spectrum of psychotic experiences may help clarify the diagnostic and etiologic ambiguity between disorders. Furthermore, it is unclear whether such impairments are specific to the patient groups or extend to unaffected first-degree relatives^37–39^. Given the shared genetic predisposition among patients and their first-degree relatives, common visual processing impairments would suggest such impairments to be an underlying risk factor rather than simply a consequence of having the disorder. Thus, by including first-degree relatives, we hoped to be able to better characterize the causality of the relationship between psychotic psychopathology and visual processing deficits.

The present study implemented a novel contrast-matched surround suppression paradigm that manipulated relative center-surround orientation (0°, 20°, 45°, 70°, or 90°) and distance (near vs. far surround conditions with inner radius at 1° and 2.5°, respectively, around a central target with a radius of .75°). Behavioral performance for each individual on this task was fit to an exponential decay function with three free parameters (*M, w*, and *o*) that represent the dependent variables of interest: the offset parameter (*o*) represents orientation-insensitive (i.e. untuned) gain-control while the magnitude (*M*), and tuning width (*w*) parameters jointly characterize orientation-dependent suppression.

Based on previous work with a similar transdiagnostic outpatient sample^7^, we hypothesized that PSZ would exhibit weakened untuned gain control (i.e., less negative offset (*o*) parameter values) and broadened tuning width of the orientation-dependent mechanisms (i.e., larger w) relative to controls. Additionally, we hypothesized that BPD, PSZrel and BPDrel would exhibit intermediate gain control and orientation-dependent suppression deficits consistent with a spectrum of psychotic psychopathology that spans conventional diagnoses reflecting that these groups share some underlying etiology with PSZ yet experience less severe phenomenological and functional disturbances. Finally, we hypothesized that PSZ would exhibit weakened suppression for both near and far surrounds suggesting a combination of impaired V1-extrinsic and V1-intrinsic mechanisms. Thus, the goals of the study were to separately characterize (1) orientation-insensitive gain-control, (2) orientation-dependent suppression magnitude and tuning width and (3) the differential functioning of these mechanisms for near and far surrounds across a spectrum of psychotic psychopathology.

## Methods

Patients were recruited from Minneapolis Veterans Affairs Health Care System (MVAHCS) outpatient clinics, community support programs for the mentally ill, and county mental health clinics. First degree relatives of PSZ and PBD were identified by research staff using a pedigree form completed through interviews with patients and were invited by mail and phone to participate in the study. Healthy controls (HC) were recruited via posted announcements at fitness centers, community libraries, the MVAHCS, and newsletters for veterans. Potential PSZ, PBD, and HC participants were excluded if they met any of the following criteria: English as a second language, age > 60 years, IQ < 70, substance dependence within the past 6 months, substance abuse within 2 weeks of testing, head injury with skull fracture or substantial loss of consciousness (i.e. loss of consciousness > 30 min), electroconvulsive therapy, amblyopia untreated before 18, epilepsy, stroke, or other neurological conditions. Additional exclusion criteria for HC were family history of major depressive disorder or a psychotic disorder (e.g. schizophrenia and/or bipolar disorder). PSZrel and PBDrel were excluded only if they had a medical condition that prevented participation.

Participants provided written informed consent before participating in the study. The study protocol was approved and monitored by the MVAHCS and the University of Minnesota Institutional Review Board. Participants were administered the Structured Clinical Interview for the DSM-IV-TR Axis-I Disorders-Patient Edition (SCID-I/P^40^), Brief Psychiatric Rating Scale, 24-item (BPRS^41^), Sensory Gating Inventory (SGI^42^), and Wechsler Adult Intelligence Scale, Third Edition (WAIS-III^43^). A minimum of two trained raters (advanced doctoral students in clinical psychology, postdoctoral researchers, or licensed psychologists) reached consensus on all diagnoses, based on the DSM-IV-TR criteria^44^. Additional participant and study information is detailed in previous publications^24,26,45^.

Group demographics for participants meeting inclusion criteria are tabulated in Table 1.

**Table 1.** Demographic and Symptom Severity Information.

| Variable | PSZ n=(31) | PBD n=(29) | HC n=(29) | PSZrel n=(28) | PBDrel n=(21) | Statistic |
| --- | --- | --- | --- | --- | --- | --- |
| Percent Female | 32 | 38 | 52 | 61 | 52 | $X^2(4)=6.269$, $p=0.18$, Cramer's $V=0.21$ |
| Age | 46.42 (8.99) | 47.41 (10.22) | 46.24 (9.63) | 47.18 (8.78) | 40.52 (11.67) | $F(4,133)=1.9$, $p=0.11$, $\eta^2=0.05$ |
| Visual Acuity (LogMAR)<sup>1</sup> | 0.14 (0.15) | 0.12 (0.12) | 0.08 (0.13) | 0.1 (0.12) | 0.04 (0.1) | $F(4,133)=2.36$, $p=0.06$, $\eta^2=0.07$ |
| Years Education | 13.32 (1.76) | 15.14 (2.29) | 15.9 (1.47) | 14.79 (2.23) | 14.86 (1.93) | $F(4,133)=6.9$, $p<.001$, $\eta^2=0.17$ |
| Parental Education (Ranking)<sup>2</sup> | 4.97 (1.43) | 4.59 (0.8) | 5.52 (1.24) | 5.07 (0.86) | 5.14 (0.91) | $F(4,130)=2.59$, $p=0.04$, $\eta^2=0.07$ |
| Estimated IQ<sup>3</sup> | 100.55 (13.68) | 101.9 (12.92) | 116.07 (13.89) | 107.36 (16.9) | 109.29 (15.86) | $F(4,133)=5.31$, $p<.001$, $\eta^2=0.14$ |
| CPZ Equivalent | 13.47 (23.05) | 2.44 (2.04) | - | 12.16 (10.99) | - |  |
| BPRS Total<sup>5</sup> | 41.74 (12.05) | 37.31 (9.73) | 26.07 (2.27) | 32.18 (8.76) | 32.1 (7.6) | $F(4,133)=13.35$, $p<.001$, $\eta^2=0.29$ |
| BPRS Positive | 10.26 (5.19) | 5.97 (1.64) | 5.03 (0.19) | 5.93 (2.71) | 5.71 (2.61) | $F(4,133)=14.35$, $p<.001$, $\eta^2=0.3$ |
| BPRS Negative | 4.03 (1.74) | 3.9 (1.5) | 3.28 (0.7) | 3.36 (1.1) | 3.67 (1.46) | $F(4,133)=1.75$, $p=0.14$, $\eta^2=0.05$ |
| BPRS Disorganized | 7.16 (2.54) | 6.45 (1.82) | 4.45 (0.78) | 5.86 (2.01) | 5.29 (1.52) | $F(4,133)=9.18$, $p<.001$, $\eta^2=0.22$ |
| SGI Total | 74.63 (36.46) | 79.96 (33.79) | 26.96 (18.2) | 50.3 (31.72) | 43.58 (32.61) | $F(4,126)=13.87$, $p<.001$, $\eta^2=0.31$ |

<sup>1</sup>LogMAR: normal is 0.0, better than normal is < 0. <sup>2</sup>Parental Education ranking: 1 is 7th grade or less, 7 is graduate or professional degree, the greater of the mother's or father's ranking is reported. <sup>3</sup>IQ was estimated using the Block Design and Vocabulary subtests of the Wechsler Adult Intelligence Scale, Third Edition. <sup>4</sup>CPZ equivalent was computed only for the subset of participants on antipsychotics: 26 PSZ, 17 PBD, and 3 PSZrel. <sup>5</sup>Brief Psychiatric Rating Scale (BPRS) score ranges: Total, 24-168; Positive, 5-35; Negative, 3-21; Disorganized, 4-28. <sup>6</sup>SGI Total range of scores: 0-180.

For each measure, potential group differences were assessed by 1-way ANOVA. Any measure that showed a potential group difference (using a conservative threshold of p < 0.1) was then tested for a relationship to the visual behavioral task using Pearson’s correlation against the parallel surround condition. The only measures that showed a correlation with surround suppression were visual acuity and estimated IQ (see supplemental Figures S3 and S5). For patients, medication (converted to CPZ equivalent^46^) was also tested for association with performance on the surround suppression task, and no association was found (r(41)=0.16, p=0.298).

All participants completed a contrast-matching task (Fig. 1; task details in legend) to assess the perceived contrast of a 1°-diameter circular grating patch presented at 3° eccentricity. Before task administration, visual acuity was measured in the same room at 2 meter viewing distance (LIGHTHOUSE Distance Visual Acuity Test, Long Island City, NY). For the task, gratings were presented in three configurations: with no surround, with an adjacent surround (at 5 different relative orientations of surrounding gradings ranging from 0° to 90°), and with a far surround (also at 5 relative orientations). Visual stimuli were displayed on an NEC 17” CRT monitor (35.1 × 26.7 cm, 1024 × 768 pixels) viewed from 61 cm. The display was calibrated to produce a linear relationship between pixel intensity value (0-255) and luminance (mean luminance 102 cd/m^2^). Stimuli were generated using PsychoPy^47^. Target stimuli were sinusoidally luminance-modulated gratings with a spatial frequency of 2 cycles/degree, masked by a circular aperture 1.5° in diameter, with edges defined by a raised cosine function. Luminance contrast of the target gratings was 80%. Targets were located at 3° eccentricity, 16° of polar angle below the horizontal meridian (so stimuli would have an asymmetric cortical representation to permit future EEG data analysis, not presented here). Targets were surrounded by a dark gray circle, 1 pixel wide, that was present throughout the experiment, to remove uncertainty about target location and to aid visual segmentation of targets from surrounds. In the near surround condition, the surround stimulus was also a sine-wave grating, 2 cpd and 80% contrast, masked by an annulus with inner radius of 1° (i.e., 0.25° gap between target and surround) and an outer radius of 2°. For the far surround condition, inner and outer radii were 2.5° and 5.0°, respectively. Because targets were centered at 3° eccentricity, surround gratings were masked (hard edge) so they did not come within 0.5° of the vertical meridian cross into the other visual hemifield. A white fixation square subtending 0.2° of visual angle was present throughout the experiment.

**Figure 1.**
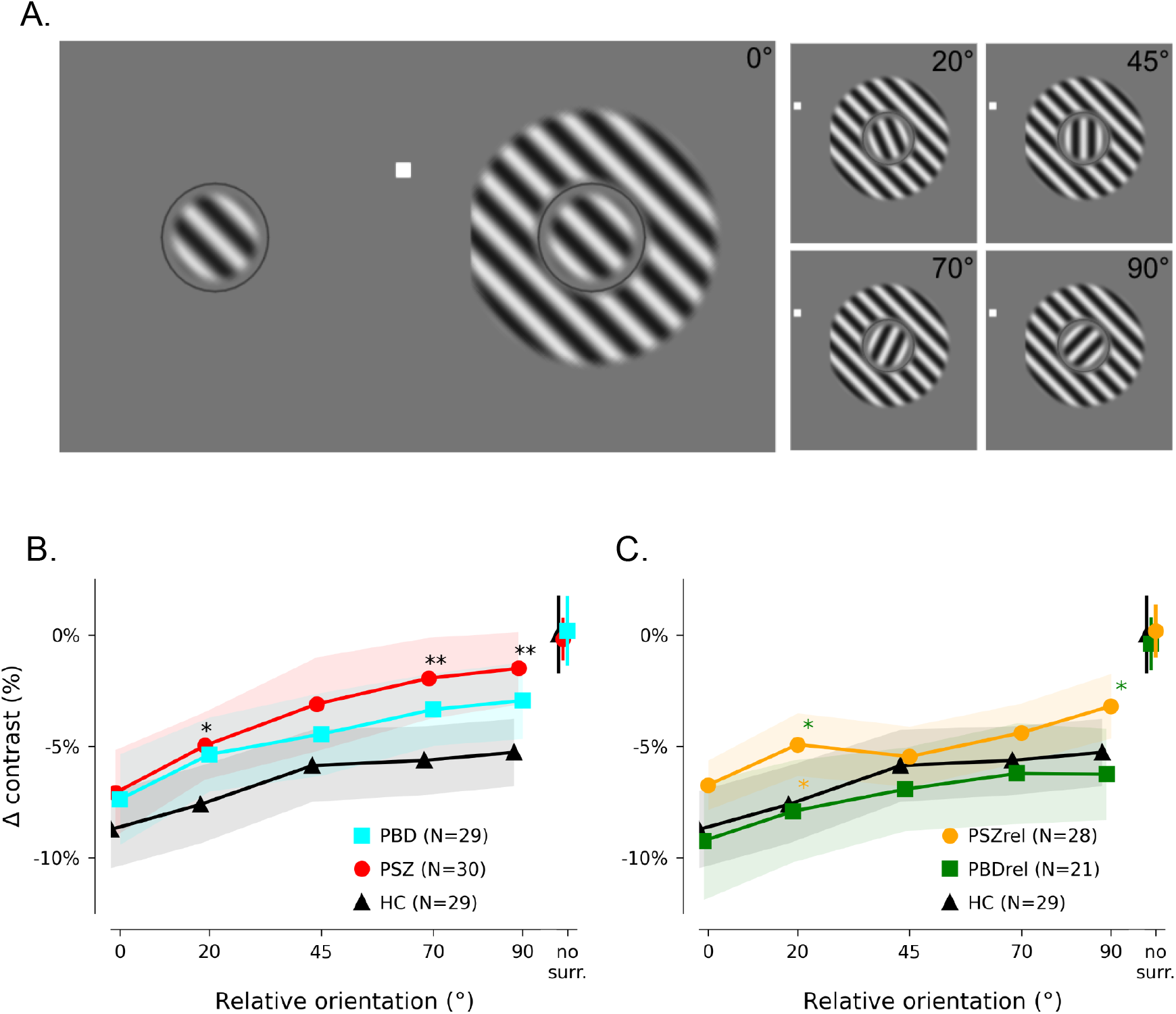
Stimulus presentation paradigm and behavioral results. Panel A: Near surround condition stimuli: the target (1.0° diameter grating with spatial frequency of 2 cycles/°) is separated by 0.25° from a surrounding annulus with an inner diameter of 1.5°. All gratings appeared simultaneously and were present for 250 msec; relative orientation of target and surround was set to one of five values, but the orientation of the target grating was randomly selected on each trial. Participants had unlimited time to respond with a button press to indicate whether the circular grating on the left or right appeared to have higher contrast. Panels B & C: Each point represents the average contrast decrement applied to the reference grating to match the perceived contrast of the target grating. Panels B & C are the same except panel B depicts the patient and control groups while panel B depicts the first-degree relative and control groups. Points with error bars indicate contrast settings for a no-surround control condition. Error bars and shaded regions represent bootstrapped 95% confidence intervals. Asterisks indicate significant differences in post-hoc t-tests (*: p < 0.05; **: p < 0.01); color of asterisk indicates group against which significant difference was measured. The same control group is presented in both plots as a reference.

A single trial consisted of the simultaneous presentation of 3 elements for 250 msec: a reference circular grating with no surround, a target circular grating, and a surrounding annulus (either near or far, at 1 of 5 possible relative orientations). The target and reference gratings were presented at a randomly selected orientation (0°, 45°, 90° or 135°) on each trial; the orientation of the target and reference was the same on a given trial. The orientation of the surround was controlled relative to the target surround. There was also a “no surround” condition in which both sides of the screen appeared identical (i.e., only the target and the reference elements appeared).

The contrast of the target stimulus was always 80%; the contrast of the reference stimulus was adjusted to achieve a match in perceived contrast. The side on which the reference stimulus was presented was randomized, so the target (plus surround) occurred on both sides of the screen with equal probability. Participants responded with a 2-button button box to indicate whether the circular patch on the left side or the right side of the screen appeared to have higher contrast.

For each condition, the contrast of the reference grating was controlled by a separate Psi staircase^48^ implemented in PsychoPy^47,49^ with the following parameters: alpha (threshold) range/precision [-40, 20]/1; beta (slope) range/precision [.1, 5]/0.05; intensity (delta-contrast for reference) range/precision [-75, 15]/1; step type: linear; delta: 0.08 (lapse rate: 4%). Each staircase converged at a point of subjective equality between the reference grating (for which contrast was varied) and target (fixed contrast) grating.

Catch trials were also embedded in the task (8% of trials were catch trials). There were 48 catch trials, evenly divided between parallel and orthogonal surrounds and randomly assigned to the near or far condition. On a catch trial, the reference contrast was fixed at 30%. On these trials, participants should have always pressed the button that indicated that the non-reference side was of higher contrast. Performance on catch trials was used to assess participant engagement in the task and compliance with task instructions.

One experimental run contained 48 staircased trials for each condition. Conditions were not blocked; trials from different conditions were mixed together because the task never changed. Participants completed 1 experimental run each, providing a single estimate of perceived contrast for each of the 11 conditions (target with no surround, 5 near surround conditions and 5 far surround conditions). The experimental run was paused 4 times so participants could rest their eyes and adjust their seat, verbally telling the experimenter when they were ready to continue.

## Analysis

Data from each participant were analyzed if they met the following criteria: accuracy on catch trials was better than 75% and their behavior indicated that perceived contrast of the target with near surround at 0° and 20° relative orientation was reduced. This last criterion was in place to eliminate participants who could not selectively attend to the central targets and instead reported the overall (target plus surround) pattern. A total of 30 datasets were discarded because they did not meet these criteria (3 PSZ, 3 BPD, 1 HC, 1 PSZrel, 0 PBDrel because of performance on catch trials and 9 PSZ, 2 PBD, 6 HC, 4 PSZrel, 1 PBDrel for reporting high contrast in parallel conditions), leaving a total of 138 datasets for analysis (31 PSZ, 29 PBD, 29 HC, 28 PSZrel, 21 PBDrel).

For each of the 11 conditions (no-surround, and 5 relative orientations for each of the near and far surround conditions), perceived contrast was calculated as the mean of the last 3 threshold estimates produced by the Psignifit staircase^50^ for that condition (excluding catch trials). The adaptive staircase failed to converge for one individual from the PSZ group for the no-surround trials which resulted in an outlier threshold value greater than 5 standard deviations from the mean for that condition only. We excluded that individual from analyses in which the no-surround threshold values were dependent variables, but included their data for all other analyses.

Following the *a priori* hypothesis that near and far surround suppression are mediated by different neural mechanisms, separate repeated measures ANCOVAs (rmANCOVAs) were performed to assess main effects of group and three of the surround conditions (0°, 90°, no-surround) and interaction between group and surround condition while controlling for visual acuity. Although we ultimately decided to include visual acuity as a covariate, such a decision is determined by theoretical perspective (e.g. ‘are visual impairments an integral component of schizophrenia or simply a confounding factor?’). Given the lack of certainty around this issue, we also report our main results *without* including acuity as a covariate (Table 2). Additionally, the choice of including fewer conditions for the rmANCOVA was driven by the fact that as the number of levels of the within-subjects factor (i.e. number of conditions) increases, the power for detecting an effect decreases. Thus, choosing fewer levels that maximize within-subject differences is preferred for the rmANCOVA.

**Table 2.**
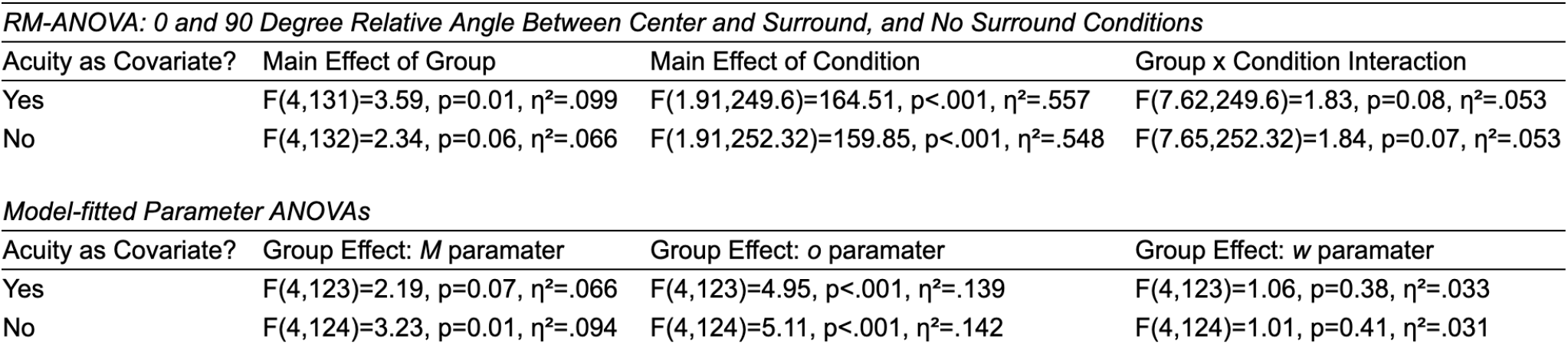
Main results with and without acuity as a covariate.

0 and 90 Degree Relative Angle Between Center and Surround, and No Surround Conditions*
| Acuity as Covariate? | Main Effect of Group | Main Effect of Condition | Group x Condition Interaction |
| --- | --- | --- | --- |
| Yes | $F(4,131)=3.59$, $p=0.01$, $\eta^2=.099$ | $F(1.91,249.6)=164.51$, $p<.001$, $\eta^2=.557$ | $F(7.62,249.6)=1.83$, $p=0.08$, $\eta^2=.053$ |
| No | $F(4,132)=2.34$, $p=0.06$, $\eta^2=.066$ | $F(1.91,252.32)=159.85$, $p<.001$, $\eta^2=.548$ | $F(7.65,252.32)=1.84$, $p=0.07$, $\eta^2=.053$ |

*Model-fitted Parameter ANOVAs*
| Acuity as Covariate? | Group Effect: *M* parameter | Group Effect: *o* parameter | Group Effect: *w* parameter |
| --- | --- | --- | --- |
| Yes | $F(4,123)=2.19$, $p=0.07$, $\eta^2=.066$ | $F(4,123)=4.95$, $p<.001$, $\eta^2=.139$ | $F(4,123)=1.06$, $p=0.38$, $\eta^2=.033$ |
| No | $F(4,124)=3.23$, $p=0.01$, $\eta^2=.094$ | $F(4,124)=5.11$, $p<.001$, $\eta^2=.142$ | $F(4,124)=1.01$, $p=0.41$, $\eta^2=.031$ |

To more fully characterize the dependence of suppression on relative orientation for each individual, we fit each participant’s contrast decrement data for the 0°, 20°, 45°, 70°, and 90° conditions to an exponential function: *P = -Me*^*-θ/w*^ *+ o*. The *o* parameter provides an estimate of the contribution of untuned gain control mechanisms, while *M* and *w* characterize the orientation-dependent mechanisms. Fitting was done in Python using scipy.optimize.curvefit non-linear least squares fitting algorithm^51^. Fits for individual participants were determined to be good if the variance of the data after subtracting the fit was lower than the variance of the raw data. By this criterion, only 9 of the 138 datasets were not well characterized by the exponential fit (1 PSZ, 3 BPD, 2 HC, 3 PSZrel, 0 PBDrel). These 9 subjects were excluded from all reported analyses in which the fit parameters were the dependent variable. Thus, it was concluded that the exponential function was an appropriate way of characterizing surround suppression behavior.

To generate hypotheses about the factors contributing to observed group differences in suppression of perceived target orientation as a function of surround orientation, a well-established divisive normalization model^52^ was adapted to simulate behavior on this dataset: *R = A*_*c*_*C*_*c*_ */ (A*_*c*_*C*_*c*_ *+ C*_*s*_*e*^− *θ*/*w*^ *+* σ). The model equation and parameters used to generate the simulated suppression tuning curves are fully detailed in the legend of Figure 4. In the original model, attention (*A*_*c*_) provides multiplicative enhancement of neuronal responses to stimuli: focal attention enhances only the target response; distributed attention enhances both target and surround responses. If this multiplicative modulatory term (*A*_*c*_) is instead used to represent the more general concept of “segmentation”, then either low acuity or broadly distributed attention (or a combination of the two) will result in stronger divisive normalization (response suppression) by the surround. Further, widely reported deficits in cortical untuned gain control associated with schizophrenia ^2,13,53^ can be simulated by decreasing the semi-saturation constant in the denominator (σ). The combination of these two terms -- weakened untuned gain control and impoverished use of segmentation cues -- recapitulates the increased sensitivity to orientation observed in the patients with schizophrenia, relative to controls. At the same time, simple reduction in scene segmentation caused by low acuity predicts the overall increase in surround suppression observed in controls. Quantitative fitting of the model to the data was not attempted; parameters were selected to illustrate how attention and suppression may interact to generate the patterns observed in the data.

## Results

The near and far surround conditions are expected to invoke different neuronal mechanisms of suppression; however, the far surround condition produced no significant modulation of perceived contrast in the present study and is therefore reported only in the Supplemental Material (Figure S1). In the near surround condition, all groups showed strong suppression of perceived contrast in the presence of a parallel surround and weaker suppression by misaligned surrounds (see Figure 1).

A repeated measures analysis of covariance was run on the three key near surround conditions (0°, 90°, and no-surround as a baseline), with Huynh-Feldt correction (ε = .991) and visual acuity as a covariate. This revealed significant main effects of group and condition (F_group_(4,131)=3.59, p=0.01, η²=.099; F_condition_(1.91,249.6)=164.51, p<.001, η²=.557; Fig. S2); however, the interaction of group and condition was not significant (F(7.62,249.6)=1.83, p=0.08, η²=.053).

Figure 2 characterizes the fitted exponential function (*P = -Me*^*-θ/w*^ *+ o*) results for each group. Group differences in the three fit parameters were assessed with one-way ANCOVAs, using acuity as a covariate. Previous work suggests that PSZ have broader orientation tuning^16,39^, and PSZ and PSZrel did tend to have fits with larger *w* parameters (broader orientation tuning), but there was not a significant difference between groups (F(4,123)=1.06, p=0.38, η²=.033). The *M* parameter showed a marginal effect of group that was not significant (F(4,123)=2.19, p=0.07, η²=.066) when acuity was entered as a covariate, but was significant without acuity as a covariate (see Table 2). The untuned gain control *o* parameter exhibited the strongest group effect (F(4,123)=4.95, p<.001, η²=.139). Follow up pairwise comparisons showed that PSZ exhibited less negative *o* parameters as compared to HCs and BPDrel (FDR corrected ps<.006).

**Figure 2.**
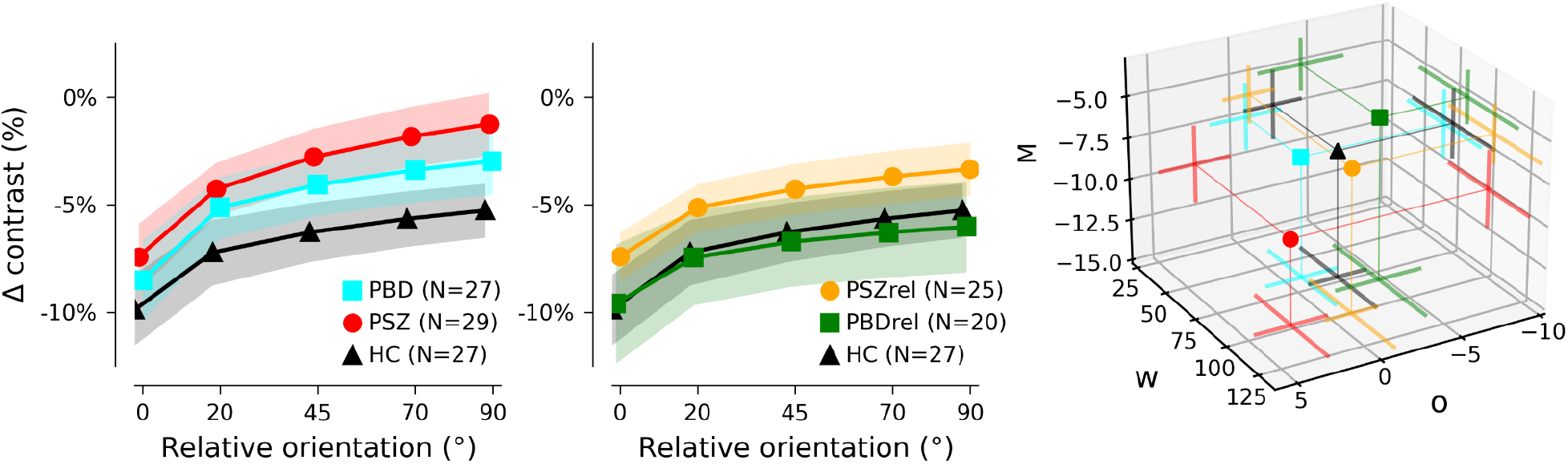
Estimation of influence of orientation on surround suppression. Each participant’s data in the near condition was fit to an exponential function describing the dependence of surround suppression on relative orientation: *P = -Me*^*-θ/w*^ *+ o*, where *M* represents modulation magnitude, *w* characterizes tuning width, and *o* estimates orientation-insensitive suppression that is present even at 90° relative orientation. Datasets for which the fit did not decrease variance are excluded from the group averages shown in this figure (3 PSZrel, 3 PBD, 1 PSZ, 2 HC, 0 PBDrel). Points indicate mean fit parameters; error bars/shading indicate 95% bootstrapped confidence intervals. Far right: average values of fit parameters describing the sensitivity to orientation (*M*), the rate of decay of suppression as a function of relative orientation (*w*), and minimum suppression (*o*). PSZ showed greater sensitivity to orientation (more negative *M*), weaker untuned gain control (less negative *o*), and – along with PSZrel – a tendency toward broader orientation tuning of suppression (higher *w*).

While there were no significant group differences in measured acuity (Table 1), acuity moderated the relationship between diagnostic group and M in a stepwise manner (F(4,119)=2.96, p=0.02, η²=.090)^54^. To illustrate this moderation effect, Figure 3 depicts each group split by LogMAR acuity at 0.1 (Snellen acuity 20/25). This visualization shows that group differences are exaggerated in participants with low acuity. For HC, reduced acuity was associated with stronger suppression in all stimulus conditions, while in PSZ, reduced acuity was associated with reduced suppression by orthogonal surrounds.

**Figure 3.**
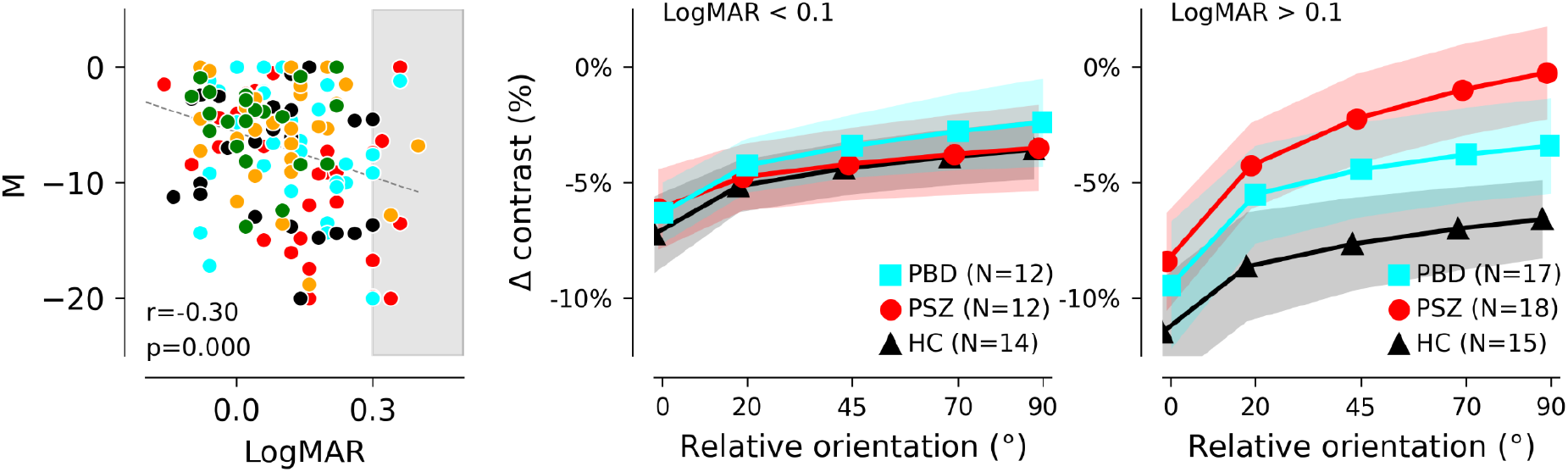
Association of perceptual suppression with acuity. Left panel: across all participants, worse acuity (higher LogMAR scores) was associated with greater modulation (*M*) of suppression between parallel and orthogonal surround conditions. Right panels: to illustrate this effect, groups were split into sub-groups of participants with acuity better than LogMAR values of 0.1 (equivalent to Snellen acuity of 20/25) and participants with acuity measured at LogMAR=0.1 or worse. Averages of individual exponential fits to suppression as a function of surround orientation are plotted here, as in Fig. 2, for controls and patient groups; for plots of average behavioral data, and data for relative groups, see Supplemental Figure S4.

Finally, we conducted exploratory correlational analyses to test whether untuned gain control as measured by the *o* parameter tracked meaningfully with individual differences in atypical sensory experiences using the SGI. To avoid the psychometric pitfalls of sum scores ^55^, we conducted a four factor oblimin-rotated EFA on the 36 item-level data and extracted ten Berge factor score estimates using the psych R package ^56,57^. The four factor solution produced the most negative BIC (Bayesian Information Criterion, BIC = -1534.58) relative to the three and five factor solution suggesting the four factor solution best balanced model parsimony and fit which is consistent with previous work^42,58^ (χ²(492)=886.05, p<.001, TLI=0.86, CFI=0.89, RMSEA=0.08). Loadings greater than .3 for each factor are presented in Supplemental Table 1. Only the scores on the third factor correlated meaningfully with the offset parameter (r(122)=0.19, p=0.037). This third factor loaded most heavily on over-inclusion items such as “I notice background noises more than other people” & “I seem to hear the smallest details of sound” though it also loaded onto a few perceptual-modulation items (e.g. “Sometimes I notice background noises more than usual”) and fatigue/stress items (e.g. “When I’m tired sounds seem amplified”).

## Discussion

While it is broadly known that schizophrenia is associated with deficits in contrast surround suppression^1,3,5,7,9,33,59^, our study finds that this deficit is not present when a high-contrast surround is parallel to a high-contrast center. This preservation of suppression in the parallel-surround condition is surprising and unique to the specific configuration we used. Although we are unaware of this effect being observed previously in the context of surround suppression, analogous findings have been reported in the context of perceptual grouping tasks in which PSZ performed similar to controls when grouping cues were strongest^60,61^. Thus it is possible that PSZ and PBD’s normative suppression for parallel-surrounds reflects a floor effect in which all groups are able to adequately process visual context when contrast is matched and center and surround orientations are aligned. This effect may have been more salient due to the matching of contrasts between center and surround relative to previously published studies in which center and surround contrasts differed.

Aside from the unexpected parallel surround finding, the reduced suppression evident for all other near surround conditions (as demonstrated by the group differences in the offset parameter, *o*) for PSZ is consistent with previous reports of weakened untuned gain control associated with PSZ ^7,13^. PBD and PSZrel exhibited intermediate deficits relative to controls and PSZ. This intermediate deficit may be indicative of a spectrum of psychotic psychopathology in which PSZ are at one end, HC are at the opposite end and PBD and PSZrel sit in the middle. Further evidence for this perspective is the fact that PBD and PSZrel also exhibited intermediate levels of atypical sensory experiences (SGI) and general psychiatric symptoms (BPRS). Indeed, individual differences in over-inclusive perceptual experiences predicted weakened untuned gain control across participants; however, this was an exploratory association and the effect size of the relationship was small such that this result needs to be replicated in an independent sample.

The dependence of contrast perception on relative orientation was moderated by acuity: surround suppression differences between patients and controls were all but eliminated when participants with lower acuity (Snellen acuity worse than 20/25) were excluded from analysis. It is noteworthy that approximately half of each of our experimental groups (patients and controls alike) had vision that was not corrected to normal (20/20) during the experiment. Our measurement of acuity was a simple Snellen eye chart at the 2-meter viewing distance that would be used for the task, with participants using any prescription lenses they had brought with them. It is possible that the relatively poor acuity across all groups was a consequence of the visual working distance being a poor match for the correction a given participant was using. To further complicate the matter, we did not observe significant differences in acuity between HC and PSZ as has been reported previously (however see ^37^). This may suggest that our convenience sample of HCs happened to have unusually poor acuity relative to the general population.

If there had been no effect of acuity in our dataset, the pattern of results shown in the full group averages (Fig. 1 and 2) might have been explained by a difficulty of the PSZ group to deploy spatial attention. While perceptual suppressive mechanisms are generally reduced for patients with schizophrenia, the same patients also experience a unique difficulty in allocating visual spatial attention or controlling attention^33^. Focal spatial attention is known to reduce surround suppression^25,62–64^. Thus, when scene segmentation cues are not strong (e.g., the parallel-surround condition, when surround and center have the same contrast), an elevation of suppression due to impairment of focal spatial attention for patients with schizophrenia could mask or counterbalance the generally observed surround suppression deficit. In other words, if impaired spatial attention had the greatest effect for stimuli with the weakest segmentation cues (in our experiment, the parallel surround), then deficits in untuned gain control for patients would emerge as relative orientation increased, and we would see the pattern shown in Figures 1 and 2: stronger modulation by orientation in patients than in controls. Previous experiments may not have detected this effect because the lower contrast of the central target relative to the surround provided a consistent, strong segmentation cue to help capture the spatial attention of all participants. Further exploration of this effect with a sample of patients with more severe symptomatology and cognitive impairment, and a rigorous assessment of spatial attention will be important for corroborating this proposed spatial attention hypothesis.

Reduced acuity could also affect task performance by altering an individual’s access to segmentation cues and thereby increasing the strength of suppression (because the V1-extrinsic mechanisms that rescue neuronal responses from suppression^18^ would be absent when boundaries are not detected). A thin black ring, always present on the screen, not only delineated the region where a participant could expect to see the target in this experiment but also formed an explicit (though subtle) boundary between the target and the surround. With poor acuity, this ring would be less visible and might even (along with the small gray gap) blend into the target and surround, removing an explicit segmentation cue and resulting in stronger suppression^19^. This explanation predicts the pattern observed for control participants: participants with low acuity showed stronger suppression of perceived contrast, compared to participants with high acuity, for all stimulus conditions. Thus, it is possible that removing this thin black ring would have led to more similar levels of suppression across levels of visual acuity in controls. However, doing so would remove the fiducial mark for the target location, increasing the spatial uncertainty and likely making it more difficult to allocate spatial attention.

Stronger perceptual suppression due to poorer acuity (and therefore reduced segmentation between center and surround) does not fully account for the relationship we observed between acuity and suppression across all participants: while low-acuity HC demonstrated greater suppression at all orientations, low-acuity PSZ showed *reduced* suppression for orthogonal surrounds and greater sensitivity to orientation. Thus, to explain the full pattern of data, we would need to posit that (1) all participants experience increased suppression when acuity reduces segmentation cues, and (2) of the PSZ group, only individuals with low acuity experience reduced untuned gain control (orientation-insensitive inhibition). Figure 4 presents a computational model that simulates the measured pattern of responses in patients and controls.

**Figure 4.**
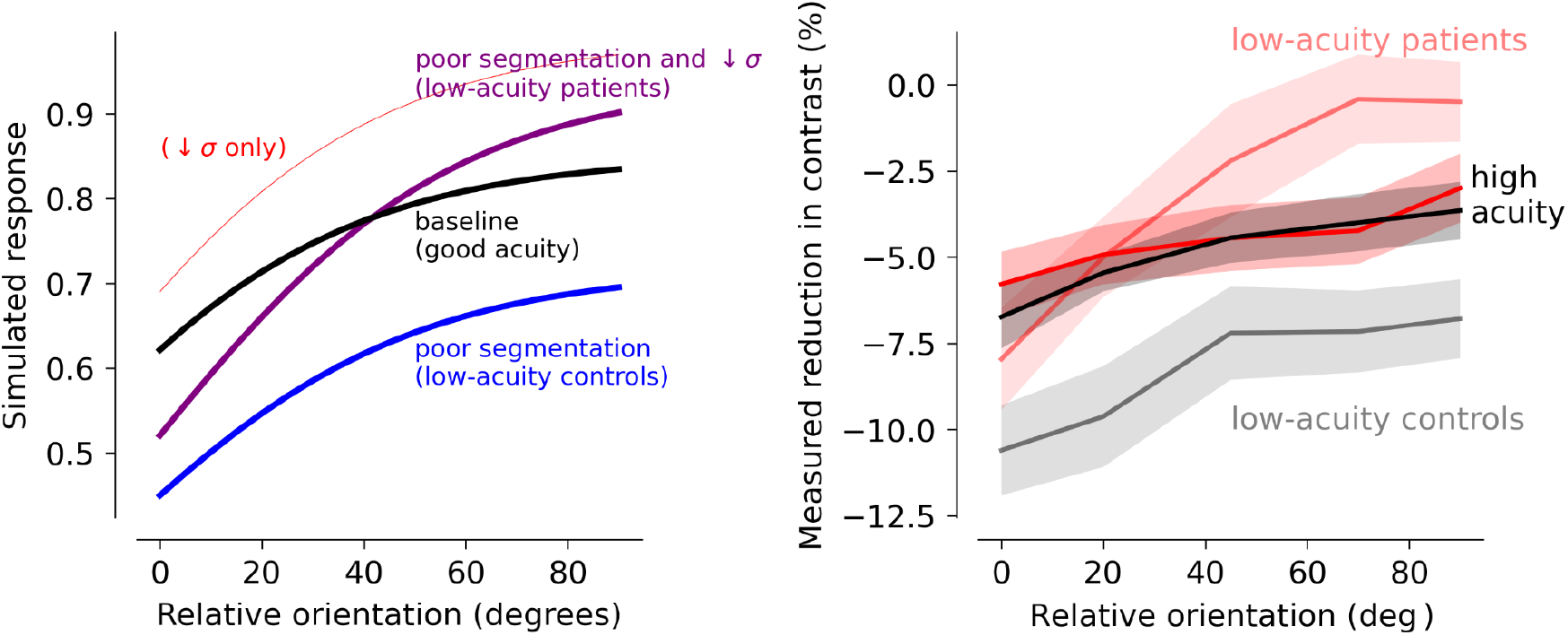
Segmentation and untuned gain control can interact to determine orientation dependence of surround suppression. Possible effects of attention and untuned gain control were simulated using a model styled after Reynolds and Heeger, 2009: *R = A*_*C*_ *C*_*C*_ **/ (***A*_*C*_ *C*_*C*_ +*C*_*S*_ *e* ^*-θ/w*^ + σ) where *R* represents the normalized response to the central target (which predicts perceived contrast), *A*_*C*_ represents amplification of the center relative to the surround, either by focal spatial attention or improved scene segmentation due to high acuity. *C*_*C*_ represents the average response to the central target, *C*_*S*_ represents the response to the surrounding stimulus, and σ is an additive constant that reflects non-specific inhibition or untuned gain control. *C*_*S*_ is modulated by an exponential term that reflects exponential dependence (*w*) of surround suppression on the relative orientation (*θ*) of the center and surround. For all simulations, *C*_*C*_ and *C*_*S*_ were held constant (0.8) to represent the equivalent contrast of the center and surround stimuli, and *w* was also fixed because the data did not provide strong evidence for group differences in the orientation tuning width of surround suppression. **Left panel:** The black line simulates a baseline condition with good use of segmentation cues or focal attention and relatively strong cortical untuned gain control (*A*_*C*_ = 2.0, σ =0.4). The blue line simulates broadly distributed attention or low acuity (*A*_*C*_ =1.0), which results in an orientation-dependent increase in the modeled strength of suppression compared to suppression during focal attention (black line). The faint red line simulates reduction of the semi-saturation constant (σ =0.1), to simulate changes in surround suppression or untuned gain control associated with schizophrenia, which causes a reduction in suppression at all relative orientations (again with some orientation dependence because the relative size of *C*_*S*_ and σ depends on the surround orientation). The purple line shows that a combination of these two factors – orientation-dependent amplification of suppression by broadly distributed attention or poor acuity and reduction of baseline suppression by a reduced semi-saturation constant – produces the pattern observed in the behavioral data from PSZ with low acuity. **Right panel**: measured behavioral data.

Further work is necessary to determine the neural mechanisms underpinning this finding that acuity moderates the relationship between clinical group and suppression. The importance of acuity and retinal health in predicting schizophrenia has received increased attention recently, and rightly so^65,66^ (and references therein). Low acuity could have several causes, ranging from inadequate optical correction (a non-neuronal source) to retinal aberrations (i.e., altered function of lateral inhibition, which sharpens boundaries) to a reduction of the cortical suppressive mechanisms necessary for accurate delineation of object boundaries (cortical acuity limits). We cannot, with the current dataset, address questions of whether acuity or retinal health is predictive of disease state in patients, although there are several known connections^66–69^. Although other researchers have found group differences in acuity with evidence that the difference is neural in origin^70^, we did not observe overall group differences in acuity in the current study. Our patient groups were outpatients with average IQ suggesting relatively normative levels of cognitive functioning. This too may have led to reductions in the magnitude of differences between controls and patients, given that links between higher IQ and stronger surround suppression have been reported among healthy adults ^71–74^. Further investigation in samples drawn from inpatients with greater cognitive impairment would be informative with respect to how illness severity impacts low-level visual deficits.

Performance of PBDrel was largely similar to performance by healthy control participants while PSZrel showed intermediate reductions in suppression at all relative orientations most similar to PBD. Within the framework of the normalization model discussed above, performance of PSZrel could arise from a decrease in the semi-saturation term (σ) in the denominator used to regulate suppression (red line, Fig. 3A). In a recent study of contour integration, we found that contour detection performance of relatives was particularly robust against suppression by flanking context^37^, to the point that their performance was superior to a control group. These findings together suggest that surround suppression might be subtly reduced by genetic liability for schizophrenia. However, a limitation of the current study is that the exclusion criteria for first degree relatives was less strict than for other groups due to the rare and valuable nature of the population. Thus it is possible that differential recruitment for these groups introduced sampling bias.

While the estimated orientation tuning of surround suppression was not significantly different between groups a non-significant effect was observed that is consistent with previous studies reporting wider orientation tuning of suppressive mechanisms in PSZ^16,39^. Broader orientation tuning associated with schizophrenia could arise either from weakened inhibitory mechanisms that refine orientation tuning of individual neurons in primary visual cortex^75,76^ or from broader tuning in higher-level grouping mechanisms^14^. V1-intrinsic and V1-extrinsic mechanisms do not work independently: a less refined V1 representation of orientation could result in a greater likelihood of grouping visual features in extrastriate cortex^77^, which in turn would result in a higher likelihood of suppression. Difficulty allocating spatial attention could readily be either caused or confounded by broader tuning of suppressive mechanisms. Additional studies exploring the physiological basis of these behavioral effects will be necessary to tease apart the separate contributions of acuity, attention, and orientation tuning in early cortical visual networks.

## Data Availability

All data produced in the present study are available upon reasonable request to the authors

## Acknowledgments

We would like to acknowledge funding from VA Merit Grant I01CX000227; NIH R01MH112583 and NIH U01MH108150. We are also deeply grateful to Andrea N. Grant for assistance with developing stimulus presentation code and for the hard work of the research assistants who helped collect data: Joseph Lupo, Haven Hafar, Abraham Van Voorhis, and Collin Teich.

## Author Contributions

Conceptualization: C.A.O., M.-P. S.; Methodology, C.A.O., S.R.S.; Formal Analysis, C.A.O., V. J. P.; Writing – Original, C.A.O.; Writing – Review & Editing, M.-P.S., S.R.S., V. J. P.; Visualization, C.A.O.; Supervision of participant enrollment and assessment, S.R.S; Project Administration and Funding Acquisition, S.R.S.

## Data and Code Availability

The published article includes all datasets generated or analyzed during this study.

## Declaration of Interests

The authors have no interests to declare.

**Supplementary Table 1.**
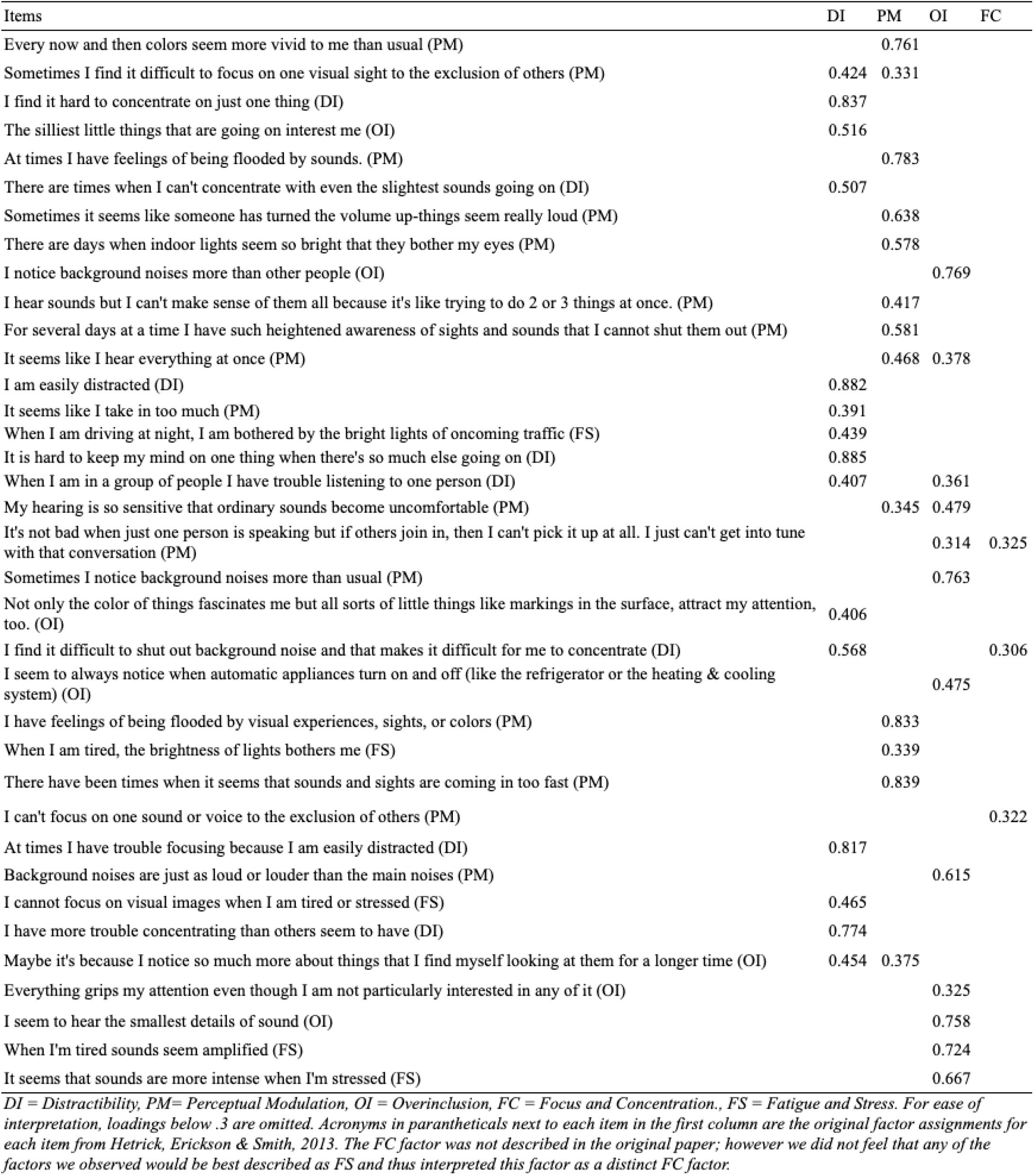
SGI Factor Loadings.

| Items | DI | PM | OI | FC |
| --- | --- | --- | --- | --- |
| Every now and then colors seem more vivid to me than usual (PM) |  | 0.761 |  |  |
| Sometimes I find it difficult to focus on one visual sight to the exclusion of others (PM) | 0.424 | 0.331 |  |  |
| I find it hard to concentrate on just one thing (DI) | 0.837 |  |  |  |
| The silliest little things that are going on interest me (OI) | 0.516 |  |  |  |
| At times I have feelings of being flooded by sounds. (PM) |  | 0.783 |  |  |
| There are times when I can't concentrate with even the slightest sounds going on (DI) | 0.507 |  |  |  |
| Sometimes it seems like someone has turned the volume up-things seem really loud (PM) |  | 0.638 |  |  |
| There are days when indoor lights seem so bright that they bother my eyes (PM) |  | 0.578 |  |  |
| I notice background noises more than other people (OI) |  |  | 0.769 |  |
| I hear sounds but I can't make sense of them all because it's like trying to do 2 or 3 things at once. (PM) |  | 0.417 |  |  |
| For several days at a time I have such heightened awareness of sights and sounds that I cannot shut them out (PM) |  | 0.581 |  |  |
| It seems like I hear everything at once (PM) |  | 0.468 | 0.378 |  |
| I am easily distracted (DI) | 0.882 |  |  |  |
| It seems like I take in too much (PM) | 0.391 |  |  |  |
| When I am driving at night, I am bothered by the bright lights of oncoming traffic (FS) | 0.439 |  |  |  |
| It is hard to keep my mind on one thing when there's so much else going on (DI) | 0.885 |  |  |  |
| When I am in a group of people I have trouble listening to one person (DI) | 0.407 |  | 0.361 |  |
| My hearing is so sensitive that ordinary sounds become uncomfortable (PM) |  | 0.345 | 0.479 |  |
| It's not bad when just one person is speaking but if others join in, then I can't pick it up at all. I just can't get into tune with that conversation (PM) |  |  | 0.314 | 0.325 |
| Sometimes I notice background noises more than usual (PM) |  |  | 0.763 |  |
| Not only the color of things fascinates me but all sorts of little things like markings in the surface, attract my attention, too. (OI) | 0.406 |  |  |  |
| I find it difficult to shut out background noise and that makes it difficult for me to concentrate (DI) | 0.568 |  |  | 0.306 |
| I seem to always notice when automatic appliances turn on and off (like the refrigerator or the heating & cooling system) (OI) |  |  | 0.475 |  |
| I have feelings of being flooded by visual experiences, sights, or colors (PM) |  | 0.833 |  |  |
| When I am tired, the brightness of lights bothers me (FS) |  | 0.339 |  |  |
| There have been times when it seems that sounds and sights are coming in too fast (PM) |  | 0.839 |  |  |
| I can't focus on one sound or voice to the exclusion of others (PM) |  |  |  | 0.322 |
| At times I have trouble focusing because I am easily distracted (DI) | 0.817 |  |  |  |
| Background noises are just as loud or louder than the main noises (PM) |  |  | 0.615 |  |
| I cannot focus on visual images when I am tired or stressed (FS) | 0.465 |  |  |  |
| I have more trouble concentrating than others seem to have (DI) | 0.774 |  |  |  |
| Maybe it's because I notice so much more about things that I find myself looking at them for a longer time (OI) | 0.454 | 0.375 |  |  |
| Everything grips my attention even though I am not particularly interested in any of it (OI) |  |  | 0.325 |  |
| I seem to hear the smallest details of sound (OI) |  |  | 0.758 |  |
| When I'm tired sounds seem amplified (FS) |  |  | 0.724 |  |
| It seems that sounds are more intense when I'm stressed (FS) |  |  | 0.667 |  |

DI = Distractibility, PM= Perceptual Modulation, OI = Overinclusion, FC = Focus and Concentration., FS = Fatigue and Stress. For ease of interpretation, loadings below .3 are omitted. Acronyms in parantheticals next to each item in the first column are the original factor assignments for each item from Hetrick, Erickson & Smith, 2013. The FC factor was not described in the original paper; however we did not feel that any of the factors we observed would be best described as FS and thus interpreted this factor as a distinct FC factor.

**Figure S1.**
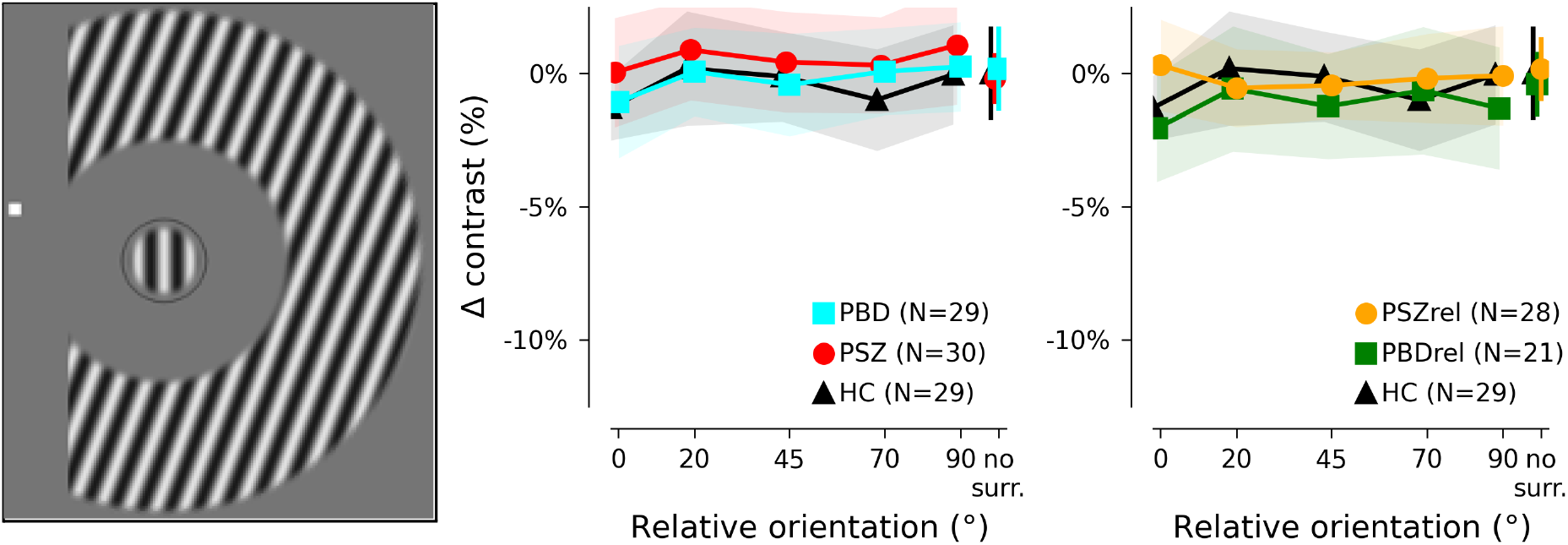
Results of far surround condition. To investigate previous reports that near and far surround stimuli produced suppression via different neuronal mechanisms, a far surround condition was included in the task design. Trial structure and analysis was identical to methods reported for the near surround condition.

**Figure S2.**
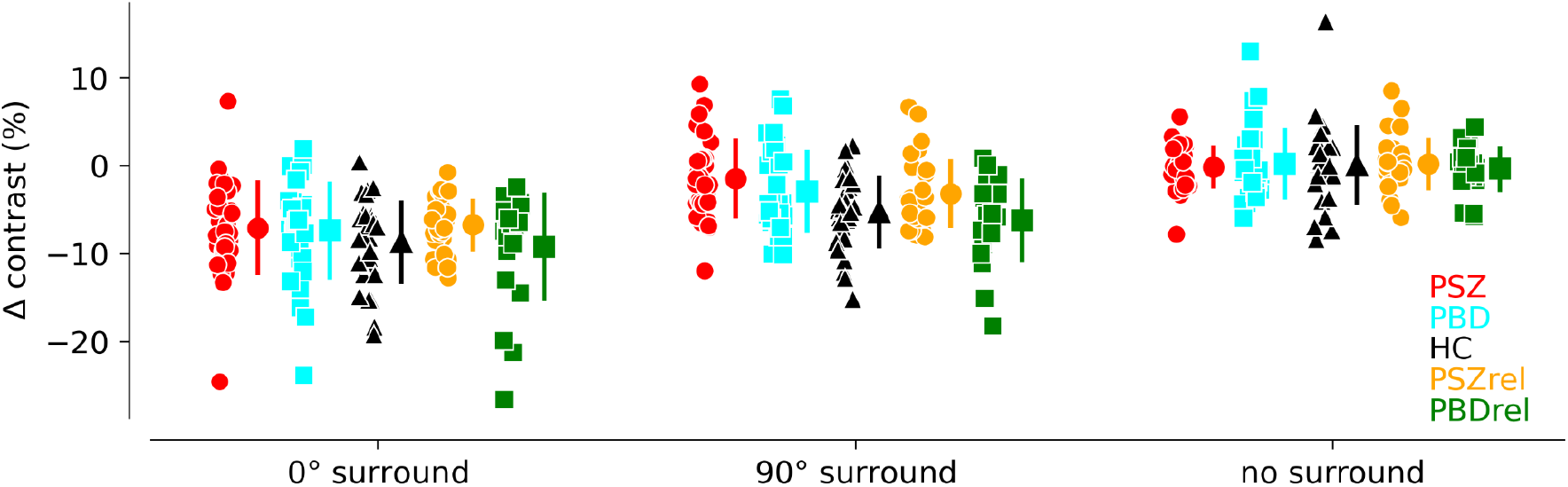
Data used for initial statistical tests of group differences in suppression. Effects were first assessed by testing performance on three conditions for each group: parallel surround, orthogonal surround, and no surround. These data points are plotted here so the reader can visually confirm the effects; small dots are behavioral values from individual participants; large dots are group means; error bars indicate standard deviation.

**Figure S3.**
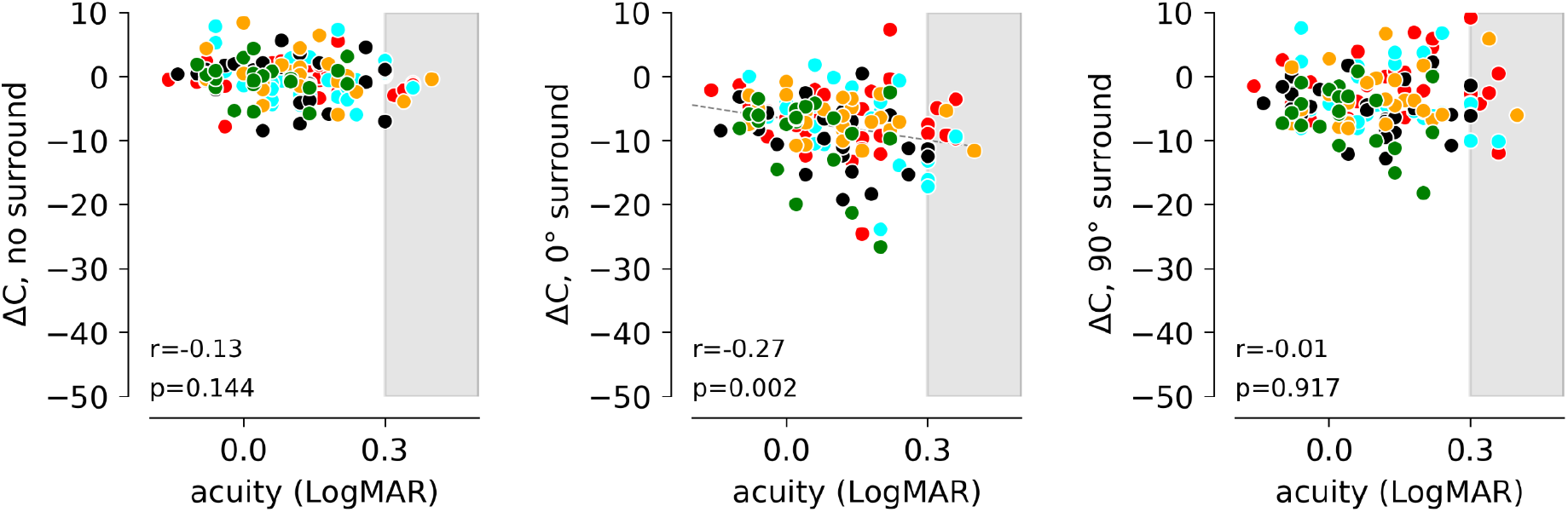
Across all groups, participants with worse acuity show stronger surround suppression. Each point represents an individual subject; mapping of color to group membership is the same as in other figures. Similar plots shown in Fig. 3 of the main manuscript show the relationship between acuity and M, the magnitude of modulation between the 0° and 90° surround conditions. Uncorrected p-values are shown.

**Figure S4.**
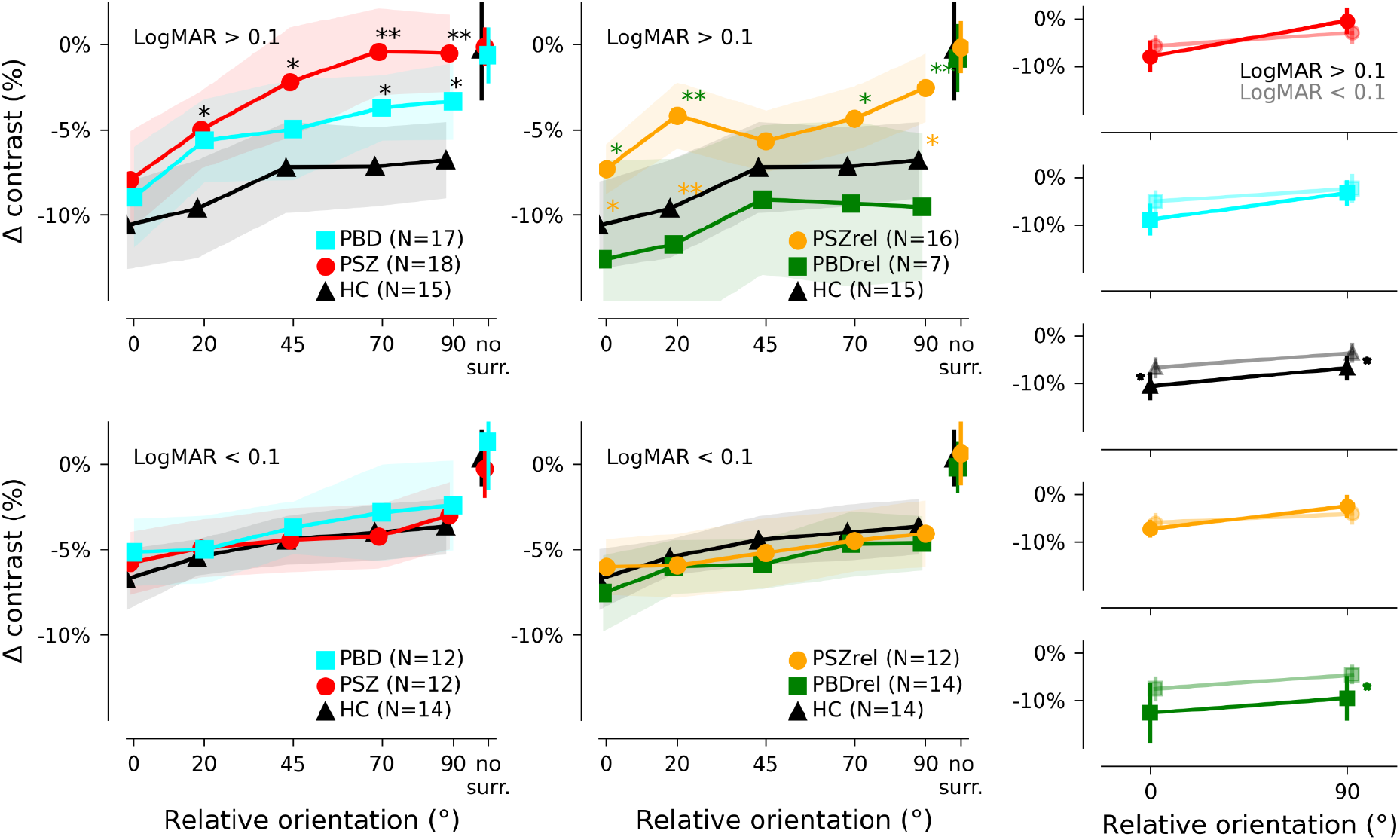
Split-half illustration of association between acuity and perceptual suppression. Figure 3 in the main manuscript shows the averaged exponential fits to individual datasets, and only for patients and controls. Here, raw behavioral data are averaged for patients, controls, and relative groups. Overall trends are the same: no differences in high-acuity sub-populations and strong differences in low-acuity groups, although variability in the smaller relative groups precludes drawing any conclusions. At far right, the extreme conditions are illustrated, illustrating the uniform shift of suppression with acuity in the HC group, and the interaction between suppression and acuity for PSZ. * indicates p < 0.05 for post-hoc paired t-tests, uncorrected. These analyses are intended for hypothesis development only.

**Figure S5.**
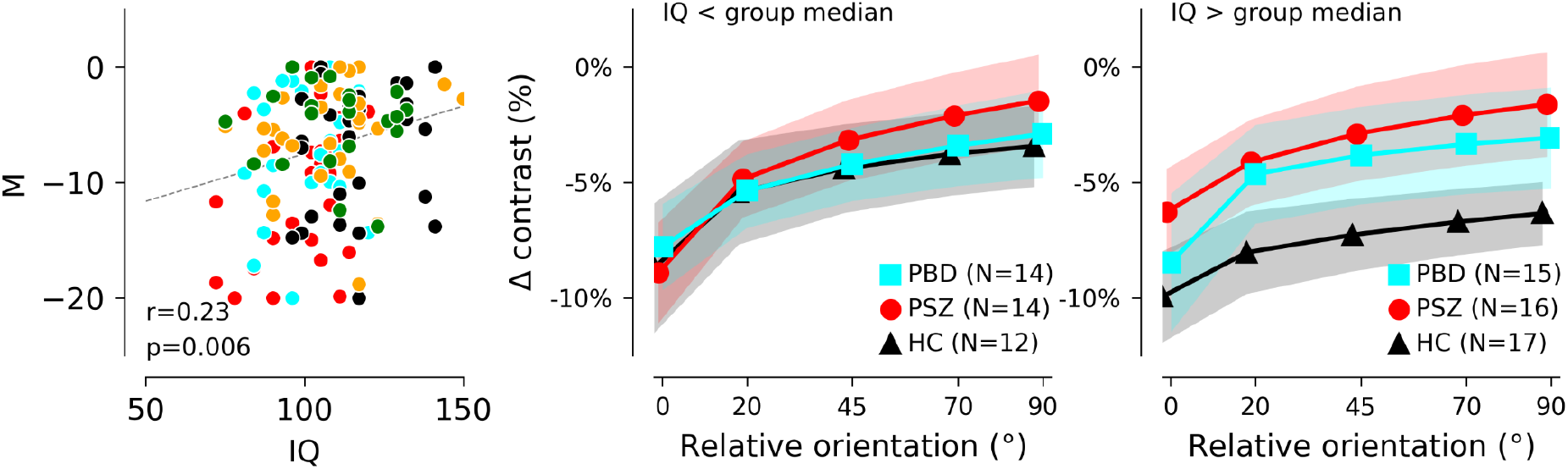
Comparisons between task performance and IQ. Because our preliminary analyses of behavioral data indicated a group difference in IQ (Table 1), and because earlier studies have reported an association between IQ and perceptual surround suppression ^32,71–74^, we also investigated the association between IQ and task performance. While, across all groups, IQ did show a relationship with *M* (left panel: higher IQ predicts less dependence of suppression on orientation, after regressing out potential effects of acuity), IQ did not significantly moderate the relationship between group and *M* (F(4,119)=0.24, p=0.91, η²=.008). Right panels: An exploratory split-halves analysis indicates that patients with higher IQ tended to have weaker surround suppression. Groups were split at the median of each group to illustrate the association between *M* and Estimated IQ. Median values for the groups shown here were 105, 105, and 114 for PSZ, PBD, and CTRL. Brief discussion: High performance on catch trials, and successful parametric manipulation of perceptual performance within individuals, indicates that this association is not simply due to a generalized cognitive deficit. While more work will be required to provide a clear answer to the question of how Estimated IQ is associated with performance on surround suppression tasks, we have considered several possible reasons for the association. Estimated IQ was measured by WAIS Vocabulary and visual Block Design (shape) tasks. The spatial imagery skills required to perform well on the block design task are housed in parietal cortex, which is also implicated as a key region for allocating spatial attention. Thus, focal spatial attention could be related to WAIS performance. An alternative mechanism by which IQ and suppression tasks might be related involves alterations in inhibition throughout the brain. In V1, altered excitation/inhibition balance is thought to produce atypical contrast surround suppression; in other brain regions, deficient inhibition may result in altered sensory and cognitive processing (e.g., reduced sensory gating, impaired mismatch detection, selective attention deficits and difficulty maintaining working memory ^23,24,78,79^). Through this mechanism, performance on surround suppression tasks would be correlated with, but not directly related to, the neural mechanisms resulting in reduced IQ.

**Figure S6.**
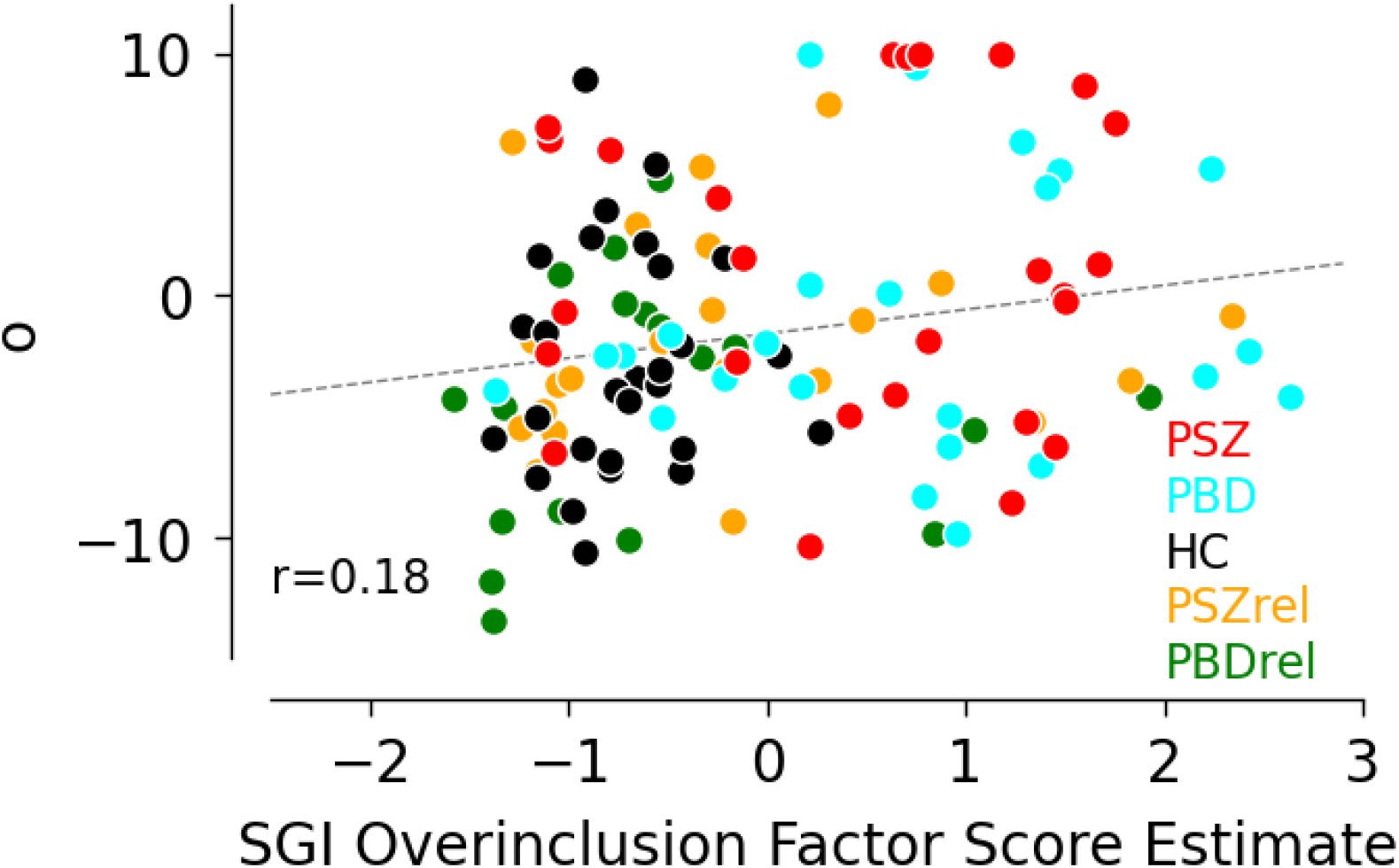
Exploratory correlation between SGI Overinclusion factor score estimates and offset parameters. Factor score estimates were derived using the ten Berge method which preserves the correlations between factors in the factor score estimates. The factor score indeterminacy (i.e. the correlation between the factor score estimates and the true factor scores) for the SGI overinclusion factor was .952.

